# Redefining Extent Of Resection After Meningioma Surgery: a Multicentre Observational Machine Learning Analysis Comparing Simpson, Radiological and Volumetric Grading

**DOI:** 10.64898/2026.05.23.26353944

**Authors:** Anand S Pandit, Matthew Deehan, Jigishaa Moudgil-Joshi, Gerda Reischer, Shalwin Mathew, Gillian Pace, Gavin Fatania, Arthur Dalton, Ramesh Nair, Harpreet Hyare, Dermot Mallon, Neil Kitchen, Hani J Marcus, Parashkev Nachev

## Abstract

**Background:** Extent of resection remains central to meningioma management, yet Simpson grading is subjective and may not reflect measurable postoperative residual disease. We compared surgeon-reported Simpson grade, report-derived radiological grading, and residual tumour volumetry across a multicentre cohort.

**Methods:** We performed a retrospective study across two tertiary neurosciences centres comprising four hospitals, including patients undergoing primary cranial meningioma resection from 2006 to 2025. Postoperative magnetic resonance imaging (MRI) reports were harmonised using weakly supervised natural language processing based on term frequency–inverse document frequency (TF–IDF) and a linear support vector machine classifier. Residual tumour volume was segmented from contrast-enhanced postoperative MRI and log-transformed. Concordance between Simpson and radiological gross-total/subtotal resection classification was assessed using absolute agreement and prevalence-adjusted bias-adjusted kappa (PABAK). Cox models assessed recurrence-free survival, with bootstrap validation and anatomical and scan-timing sensitivity analyses.

**Results:** Among 912 patients, recurrence or residual progression occurred in 281. Surgical–radiological agreement was substantial but imperfect (absolute agreement 74%; PABAK 0.61), with lower agreement in skull-base and parafalcine–parasagittal tumours. In adjusted models, recurrence hazard increased with Simpson grade (hazard ratio 1.54, 95% confidence interval 1.37–1.72), radiological grade (1.92, 1.68–2.20), and log-transformed residual volume (1.20, 1.16–1.24; all p<0.0005). Optimism-corrected concordance increased from Simpson grade to radiological grade and log-volumetry (0.692, 0.733, and 0.748), with this ranking preserved across sensitivity analyses.

**Conclusions:** Imaging-based postoperative residual disease measures outperformed Simpson grade. TF– IDF-assisted report-derived grading provides a scalable bridge to volumetry, while quantitative residual volume offers the strongest prognostic representation.

**Importance of Study:** Extent of resection remains central to meningioma management, yet clinical practice still relies on Simpson grading, a subjective intra-operative scale developed before modern MRI and volumetric assessment. This multicentre study directly compares surgeon-reported Simpson grade, machine learning-assisted radiological grading derived from routine postoperative MRI reports, and semi-automated residual tumour volumetry against recurrence outcomes. We show that operative and radiological classifications are imperfectly concordant, particularly in skull-base and parasagittal– parafalcine tumours. Increasingly objective measures provided progressively stronger prognostic information: volumetry performed best, while report-derived radiological grading outperformed Simpson grade and offered a scalable intermediate framework using routine data. The study demonstrates a novel use of routine radiology reporting and weakly supervised machine learning to harmonise residual-disease terminology across centres. These robust findings support structured radiological and volumetric assessment of the postoperative meningioma state and provide a foundation for future automated segmentation, recurrence-risk modelling, and trial stratification.

## Introduction

Meningiomas are the most common primary intracranial tumours in adults, comprising almost 40 % of all brain neoplasms ^1^. Although many are biologically indolent, they represent a substantial proportion of operative neuro-oncology practice because symptomatic, enlarging, or anatomically threatening lesions frequently require resection, with the goal of achieving maximal safe removal to minimise recurrence. The extent of resection (EOR) or residual tumour volume is a key modifiable predictor of recurrence-free and overall survival ^2^. Accurate estimation of residual disease is crucial for post-operative decision-making, including surveillance intervals and adjuvant radiotherapy selection ^3^.

Since its introduction in 1957, the Simpson grading system has remained the most widely applied intra-operative measure of EOR ^4^. Its five-point hierarchy links progressively more extensive resections with lower recurrence risk. Yet despite its historic and pervasive influence ^5^, the Simpson system has several limitations in contemporary practice. It relies on the surgeon’s subjective intra-operative impression, was designed before the advent of microsurgical and imaging technologies, and does not account for the quantitative residual tumour volume.

Post-operative MRI potentially offers a more objective assessment of residual enhancing disease, but radiological interpretation can also be imperfect, particularly in the early phase after surgery. Reactive dural enhancement, surgical and blood products, and anatomical complexity can obscure the boundary between true residual tumour and treatment effect on MRI ^6^, yet asymmetric dural enhancement and thickening have been found to be associated with tumour invasion ^7^. Previous studies have shown only moderate agreement between Simpson grade and early post-operative MRI, with clinically relevant discordance in a substantial minority of cases (7,8). Voxel-wise segmentation provides the most direct quantitative estimate of residual tumour burden and has shown promise as a predictor of recurrence ^2^, but remains labour-intensive and inconsistently implemented in routine practice. A standardised radiological framework that is reproducible, scalable, and outcome-relevant could therefore fill an important translational gap between subjective operative or radiological grading and full volumetric analysis. However, no large multicentre study has systematically assessed concordance between these methods or determined which provides the most informative prediction of recurrence.

We therefore undertook a multicentre observational study to reappraise the definition of EOR in meningioma surgery. The primary objective was to evaluate concordance between intra-operative Simpson grading and post-operative radiological grading in the classification of gross total resection, in the absence of a ground truth. The secondary objective was to determine which metric: Simpson grade, radiological grade, or residual tumour volume - best predicts recurrence over time. To our knowledge, this is the first study to integrate operative assessment, post-operative imaging, and volumetric analysis, alongside machine learning (ML)-assisted radiology report harmonisation, to develop a preliminary iteration of a scalable and reproducible framework that objectively assesses resection extent following meningioma surgery.

## Methods

This retrospective multicentre observational study was conducted in accordance with the STROBE guidelines ^8^. The study was approved by our local institutional research and development office (Study ID: 20241023, R&D Ref ID: 179421) and quality and safety committee (Reference: 1202) which confirmed the project met criteria for secondary analysis of routinely collected data.

### Patients and centres

Data were obtained from two tertiary neurosciences centres in London (each comprising two hospitals) with high-volume meningioma practices. A retrospective review of the institutional meningioma resection databases was performed to identify eligible patients who underwent primary surgical resection of a histologically confirmed cranial meningioma between 2006 and 2025. Other inclusion criteria were: (1) documented surgeon-estimated EOR (Simpson grade), (2) availability of at least a contrast-enhanced T1-weighted MRI performed within six months of surgery, and (3) accessible digital imaging suitable for volumetric analysis.

MRI scans were retrieved from institutional Picture Archiving and Communication Systems (PACS) in DICOM format and converted to anonymised NIfTI files using MRIcron (v1.0). Header metadata were stripped to remove patient identifiable data. When multiple post-operative MRI scans were available within six months of surgery, the later scan was selected for volumetric segmentation and analysis, as it was more likely to represent a stable post-operative baseline (Dolinskas and Simeone, 1998).

### Tumour characteristics

For statistical analyses, tumour location was categorised into the following anatomical groups: convexity, parafalcine–parasagittal, skull base which included cerebellopontine angle, anterior midline, sella, parasella, sphenoid wing, and foramen magnum, and ‘other’ locations which comprised intraventricular, pineal, and intraosseous locations. Tumour location categories were not mutually exclusive.

### Grading

#### Simpson Grade

When the Simpson grade was not explicitly stated, it was inferred by matching key descriptive phrases within the operative notes (e.g. dural coagulation, bone removal, or subtotal resection) to the corresponding Simpson category. All inferred grades were reviewed and confirmed by consensus. Grading followed the original five-tier classification (Simpson, 1957).

#### ML-assisted radiological grading

Radiological grading was assigned from a post-operative MRI reported by a board-certified neuroradiologist using a weakly supervised natural language processing workflow ^9^ designed primarily to support development and harmonisation of the reporting lexicon rather than to serve as a fully autonomous classifier. Free-text reports were extracted and transformed into term frequency–inverse document frequency (TF–IDF) representations using word- and character-level n-grams. A linear support vector machine (LinearSVC) model was trained on a manually labelled subset of reports mapped to the standardised radiological EOR grading system (Grades I–V; Table 1). Model outputs, feature weights, and recurrent misclassifications were then reviewed iteratively to identify adjacent, synonymous, and centre-specific expressions used to describe similar residual-disease states. These data-driven synonym candidates informed iterative refinement of the grading lexicon. Gross-total resection (GTR) grades were defined as reported complete resection either without residual or adjacent dural enhancement (Grade I) or with this finding (Grade II) ^7^. Subtotal resection (STR) grades were classified according to the reported extent of residual disease, ranging from small focal or nodular enhancement at the cavity margin (Grade III) ^10^ to a clear subtotal resection or more substantial residual tumour (Grade IV), and a largely unchanged tumour bulk following decompression- or biopsy-only procedures (Grade V). Blank reports, indeterminate reports, and reports containing terminology suggestive of infection or reactive post-operative change were assigned NA and excluded from further analysis. Additional details are provided in the Supplementary Methods. One in five reports were re-assessed for fidelity by an independent neuroradiologist, with concordance regarding residual presence or absence above 98%.

**Table 1.** Radiological grading lexicon for post-operative meningioma MRI reports following five iterations. Grades 1–2 approximately represent GTR, whereas Grades 3–5 represent STR, and NA denotes indeterminate post-operative appearances. Representative synonym terms used in the ML-assisted harmonisation process and the corresponding exclusion rules are shown for each grade.

| Grade | Definition | ML synonym lexicon | Exclusions and notes | GTR/STR |
| --- | --- | --- | --- | --- |
| I | Complete resection, no adjacent dural enhancement | "No residual tumour", "complete resection", "gross total resection", "CSF cavity only", "no enhancing tissue" | Must not mention dural enhancement, nodule, or bulk. | GTR |
| II | Complete resection with adjacent dural enhancement or thickening only | "Dural thickening", "linear dural enhancement", "enhancing dural tail", "thickened dura", "enhancing dural edge" | If nodular or globular enhancement present → Grade 3<br>if multi-compartmental or plaque like residual → Grade 4 |  |
| III | Single focus of enhancement, nodule or nodular residual at surgical margin or within cavity | "Single enhancing nodule", "tiny/small/minimal/minute residual", "punctate enhancing focus" | Exclude if residual described as multifocal, subtotal, bulky, solid or if any dimension diameter given is $\geq 10\text{mm}$ → Grade 4. | STR |
| IV | Subtotal / partial resection or multifocal residual or enlarging residual | "Subtotal resection", "partial debulk", "residual bulk", "multifocal enhancing nodules", "large remnant", "solid residual", "significant remnant", "clear residual", "progressive/enlarging/growing/increasing nodule" |  |  |
| V | Decompression or biopsy only | "Biopsy", "decompression", "no tumour removed", "bulk unchanged", "minimal debulk", "tumour unchanged" | Reserved for limited or non-resective intent. |  |
| NA | Indeterminate | "Empyema", "abscess", "meningitis", "infective/reactive enhancement", "cannot exclude residual" | Infection override: classify as NA unless biopsy-only. |  |

#### Volumetric segmentation

Volumetric segmentation was performed using ITK-SNAP (v3.8)^11^, following previously published semi-automated methods ^12,13^. Segmentation was performed blinded to both operative and outcome data and verified by a board-certified neuroradiologist specialising in meningioma and skull base lesions. If no measurable residual tumour was identified, the residual volume was recorded as 0 cm^3^. Contrast-enhanced T1-weighted MRI was used in all cases. For non-isometric scans, if available, segmentation was performed in the two most informative orthogonal planes (axial, sagittal, or coronal), and a mean volume was taken. Supporting sequences—including T2-weighted, T2-FLAIR, and pre-operative contrast-enhanced T1-weighted images, if available, were reviewed to distinguish residual tumour from post-operative changes.

### Data analysis

Study size was determined pragmatically from all eligible cases in the resection databases. Tumour recurrence and residual progression were defined radiologically. The date of recurrence was taken as the midpoint between the last stable scan and the first showing definite growth or new enhancement. Concordance between surgical and radiological gross/subtotal resection (GTR/STR) grading was assessed using absolute agreement and prevalence-adjusted, bias-adjusted kappa (PABAK). PABAK is a concordance measure derived from the observed proportion agreement and adjusts for the effects of category prevalence imbalance, thus providing a more stable and interpretable estimate ^14^.

Cox proportional hazards models were applied to evaluate the predictive value of Simpson grade, radiological grade, and residual tumour volume for recurrence-free survival. The highest radiological grade was chosen if more than one scan was present earlier than 6 months, and this scan was also used for segmentation. Hazard ratios (HRs) with 95% confidence intervals (CI) were reported, and *p*<0.05 was considered statistically significant. Adjustment was made for established prognostic factors, including age, sex, WHO grade, and use of adjuvant radiotherapy (Supplementary Methods). Model assumptions were assessed for each Cox proportional hazards model with the proportional hazards assumption evaluated using Schoenfeld residuals. The predictive performance of the Cox proportional hazard models was assessed via Harrell’s Concordance Index (C-index) which offers a numerical representation of how well the model attributes higher risk to those that experienced the event ^15^.

Four additional analyses were used to assess model validation, calibration and robustness. Firstly, internal validation was performed using bootstrap resampling (n=500) to estimate corrected model discrimination and fit. Two sensitivity analyses, using the same bootstrap implementation were performed to determine the subgroup effects of tumour location and scan timing after surgery on model performance. Finally, a nested model comparison was performed to clarify if there was incremental value in combining models together.

## Results

### Patient Characteristics

912 patients were identified meeting the inclusion criteria across both neurosurgical centres, of which 653 (72%) were female (Table 2). Seven patients were later excluded who had an artefactual MRI scan with no other usable scan within the six-month period. There were no significant site-based differences in age, sex, proportions of tumours by WHO grade, Simpson grade, recurrence rate or mean residual volume. There was a significant difference in gross tumour location by site, primarily driven by a relatively greater proportion of skull-base tumours performed at one of the centres. Fifty-three patients (6%) had a tumour which encompassed more than one location.

**Table 2.**
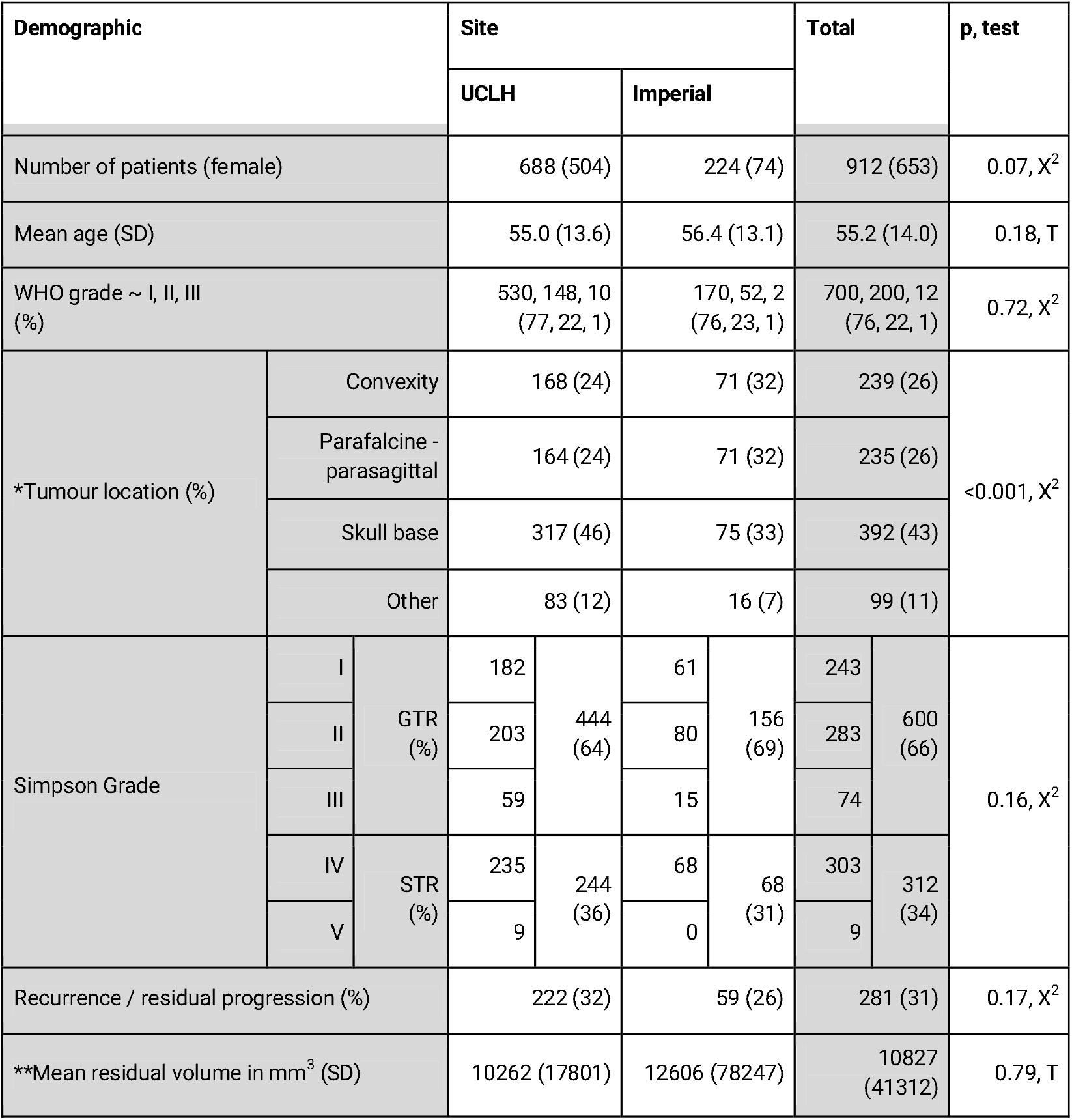
Demographic and tumour characteristics of study patients. *Tumour locations are not mutually exclusive. ** Mean volume of radiologically identified residuals. GTR = gross total resection; STR = sub-total resection

### Surgical-radiological concordance

Following ML processing and radiology report grading, concordance between the radiology-based grading scale and Simpson grade was assessed across the main tumour locations (Figure 1). Overall, agreement was substantial (PABAK 0.61; absolute agreement 74%), but varied by anatomical site. Concordance was strongest for convexity meningiomas (PABAK 0.62; abs. agreement 81%) and for other sites, including intraventricular and intraosseous tumours (PABAK 0.58; abs. agreement 79%). By contrast, agreement was lower at the skull base (PABAK 0.49; abs. agreement 75%) and lowest in the parafalcine– parasagittal region, where there was only fair concordance (PABAK = 0.32; abs. agreement 66%).

**Figure 1.**
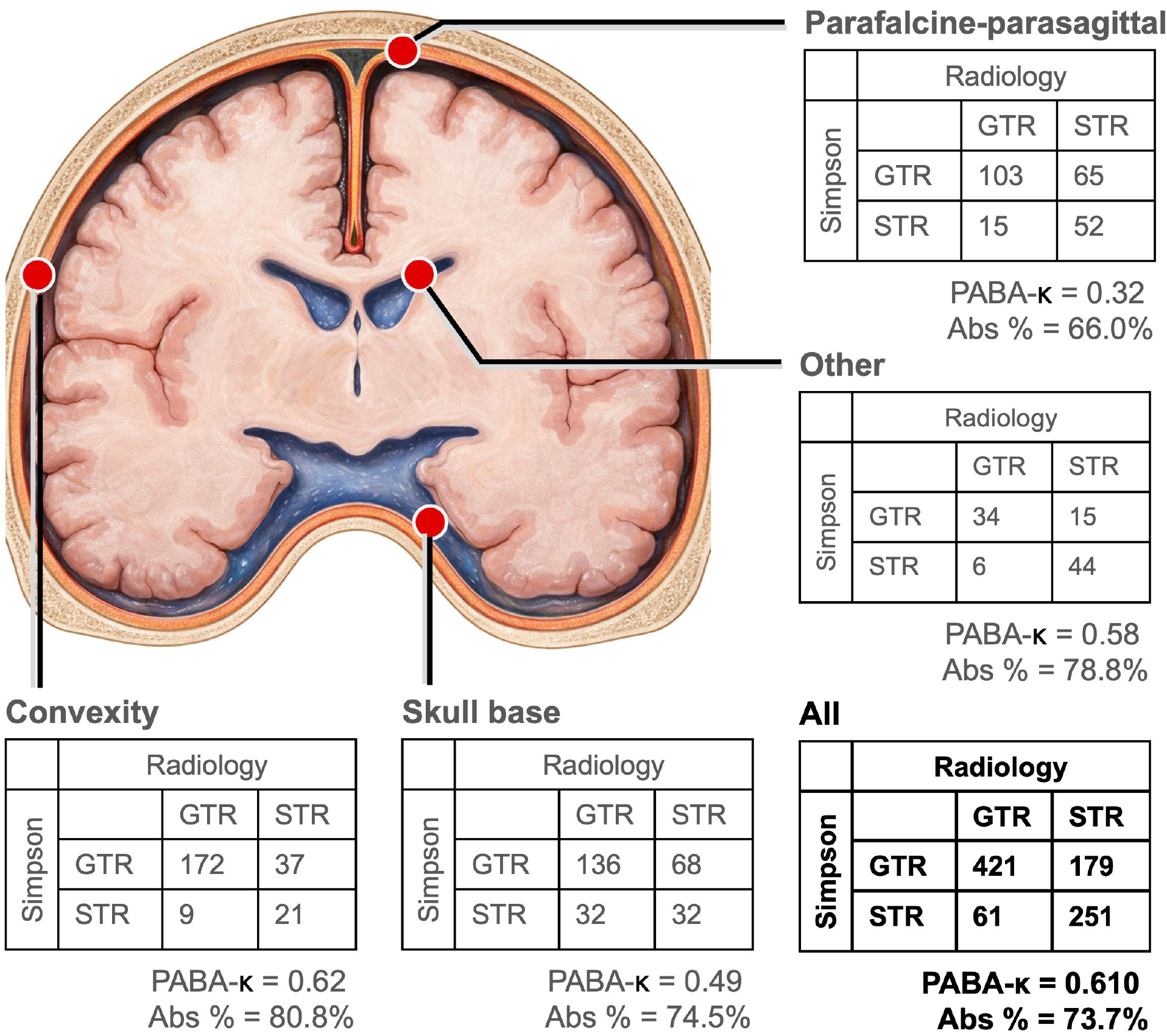
Surgical–radiological concordance for gross total resection (GTR) versus subtotal resection (STR) across anatomical resection sites. Abs% = absolute percentage agreement; PABAK = prevalence-adjusted bias-adjusted kappa. Image produced and modified using BioRender, Microsoft PowerPoint and ChatGPT5.4.

### Survival analysis

Kaplan–Meier analysis (Figure 2) demonstrated clear stratification of recurrence-free survival by both Simpson grade and ML-assisted radiological grade (log-rank, Simpson: Χ^2^ = 74.7, p <0.0001; Radiological: Χ^2^ = 124.0, p <0.0001). In each system, lower grades were generally associated with more favourable recurrence-free survival. The radiological classification showed stronger short and medium-term separation of the Kaplan–Meier curves than Simpson grade, with a larger log-rank test statistic in the early follow-up window (restricted log-rank to 5 years, Χ^2^ = 125.2 vs 72.5). Interpretation of grade V in both grading systems was limited by low patient numbers.

**Figure 2.**
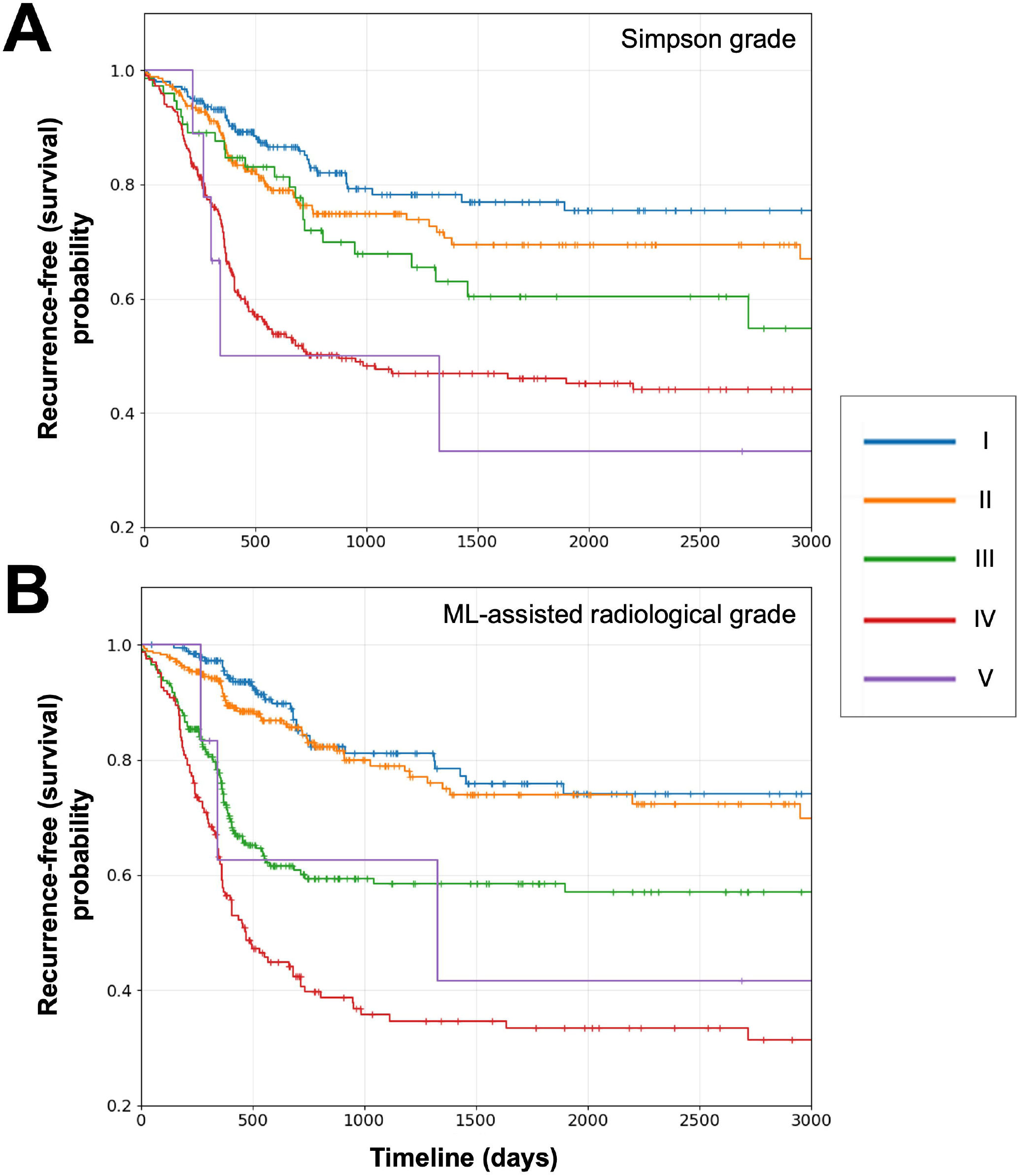
Kaplan–Meier estimates of recurrence-free survival according to (A) Simpson grade and (B) ML-assisted radiological grade. Legend box (right) showing grade colour coding.

### Volumetrics and recurrence prediction

Median follow-up was 915 days (2.5 years) by reverse Kaplan–Meier analysis (IQR 365–1343 days), with a maximum follow-up of 6884 days (18.8 years). Because residual tumour volume was markedly right-skewed, it was log-transformed before analysis. Parallel Cox proportional hazards models were then fitted, each incorporating a single index measure of EOR—Simpson grade, ML-assisted radiological grade, or log-transformed residual volume—to examine associations with time to recurrence (Table 2).

In univariable analysis, all three EOR measures were significantly associated with recurrence. Discrimination improved progressively from Simpson grade (HR 1.52, 95% CI 1.37–1.68; C-index 0.645) to ML-assisted radiological grade (HR 1.80, 95% CI 1.60–2.01; C-index 0.691), and was highest for log-transformed residual tumour volume (HR 1.18 per log-unit increase, 95% CI 1.15–1.22; C-index 0.707). This pattern suggested that increasingly objective and quantitative representations of residual disease provided greater prognostic information than Simpson grading alone.

This ranking persisted after multivariable adjustment. The log-volumetric model showed the best overall performance, with the highest discrimination and best fit (C-index 0.76; AIC 3347), followed by the ML-assisted radiology model (C-index 0.74; AIC 3369), and then the Simpson model (C-index 0.70; AIC 3340). In the adjusted models, all three EOR measures were independently associated with recurrence. Each one-grade increase in Simpson grade was associated with a 54% higher recurrence hazard (HR 1.54, 95% CI 1.37–1.72; p<0.0005), whereas each one-grade increase in ML-enhanced radiological grade was associated with a 92% higher recurrence hazard (HR 1.92, 95% CI 1.68–2.20; p<0.0005). In the volumetric model, each 1-unit increase in log-transformed residual tumour volume was associated with a 20% higher recurrence hazard (HR 1.20, 95% CI 1.16–1.24; p<0.0005), equivalent to an approximately 13–14% increase in recurrence hazard per doubling of residual volume.

Time-dependent AUC analysis demonstrated consistently better discrimination for the log-volumetric model than for ML-assisted radiology or Simpson grade throughout follow-up (Supplementary Figure 1). Mean AUC was highest for the log-volumetric model (0.790), followed by ML-assisted radiology (0.781), with Simpson grade performing least well (0.727). All three models showed strongest discrimination early after surgery, followed by a modest decline and subsequent plateau over mid-to late follow-up. Taken together, these findings indicate that volumetric residual tumour burden carries the strongest prognostic signal, with ML-assisted radiological grading outperforming conventional Simpson grading.

Among the adjustment covariates, increasing age was associated with a small reduction in recurrence hazard across all models. WHO grade remained a strong independent predictor, with grade II tumours conferring an approximately two-fold higher recurrence hazard than grade I and grade III tumours conferring the highest risk. By contrast, sex, site, tumour location, and radiotherapy were not independently associated with recurrence. Post-operative surveillance status and second-stage surgery were positively associated with recurrence, most likely reflecting greater residual disease burden or overall disease complexity rather than direct causal effects.

To support interpretation and practical application of these findings, formula-based examples and an accompanying Microsoft Excel calculator were developed to demonstrate model use at the point of analysis.

### Validation, calibration and sensitivity analyses

In a head-to-head comparison using the same patient dataset (Table 3), the log-volumetric model retained the highest corrected discrimination (C-index 0.747), followed by ML-assisted radiology (0.736) and Simpson grade (0.689). It also showed the best model fit, with the lowest AIC. The small reduction of these values after correction suggests limited effects of overfitting. Calibration at 3 and 5 years was acceptable for all three models (Supplementary Figure 1), but was visually strongest for the log-volumetric model, which showed the closest agreement between predicted and observed recurrence probabilities across the risk spectrum; ML-assisted radiology was intermediate, whereas Simpson grade showed the greatest deviation from ideal calibration.

**Table 3.** Cox proportional hazard performance across extent of resection models. AIC = Akaike Information Criterion; C-index = Harrell’s Concordance Index; CI = confidence interval; EOR = extent of resection

|  |  | Simpson |  | ML-assisted radiology |  | Log-volumetric |  |
| --- | --- | --- | --- | --- | --- | --- | --- |
| Univariate |  |  |  |  |  |  |  |
|  |  | HR (CI) | p | HR (CI) | p | HR (CI) | p |
|  |  | 1.52 (1.38–1.68) | <0.0001 | 1.80 (1.60–2.01) | <0.0001 | 1.19 (1.15–1.22) | p < 0.0001 |
|  |  | C-index = 0.645 |  | C-index = 0.691 |  | C-index = 0.701 |  |
| Multivariate |  |  |  |  |  |  |  |
|  |  | HR (CI) | p | HR (CI) | p | HR (CI) | p |
| Predictor | EOR Grade | 1.54 (1.37 - 1.72) | <0.0005 | 1.92 (1.63–2.16) | <0.0005 | 1.20 (1.16–1.24) | <0.0005 |
| Demographics | Age | 0.99 (0.98–1.00) | 0.03 | 0.99 (0.98–1.00) | 0.01 | 0.99 (0.98–1.00) | 0.01 |
|  | Female | 0.92 (0.71–1.20) | 0.55 | 1.02 (0.78–1.32) | 0.91 | 1.07 (0.83–1.39) | 0.60 |
|  | Site | 0.98 (0.72–1.33) | 0.86 | 0.99 (0.72–1.34) | 0.92 | 0.88 (0.65–1.20) | 0.42 |
| Tumour location | Convexity | 1.26 (0.72–2.19) | 0.41 | 1.20 (0.69–2.10) | 0.51 | 1.28 (0.79–2.10) | 0.32 |
|  | Parafalcine-parasagittal | 1.62 (0.92–2.73) | 0.09 | 1.67 (0.94–2.95) | 0.08 | 1.56 (0.95–2.55) | 0.08 |
|  | Skull base | 1.59 (0.92–2.73) | 0.10 | 1.51 (0.87–2.61) | 0.14 | 1.43 (0.89–2.30) | 0.14 |
|  | Other | 1.62 (1.03–2.56) | 0.04 | 1.57 (0.99–2.48) | 0.06 | 1.35 (0.88–2.08) | 0.17 |
| WHO | I | reference |  |  |  |  |  |
|  | II | 2.18 (1.65–2.87) | <0.0005 | 2.34 (1.77–3.09) | <0.0005 | 2.15 (1.63–2.83) | <0.0005 |
|  | III | 5.26 (2.25–12.30) | <0.0005 | 4.10 (1.76–9.54) | 0.001 | 3.89 (1.67–9.05) | 0.002 |
| Post-operative | Surveillance | 3.07 (1.29–7.30) | 0.01 | 2.83 (1.19–6.73) | 0.02 | 2.03 (0.97–4.27) | 0.02 |
|  | Radiotherapy | 1.72 (0.78–3.81) | 0.19 | 1.36 (0.61–3.03) | 0.45 | 0.98 (0.49–1.97) | 0.95 |
|  | 2nd stage surgery | 2.81 (1.29–6.09) | 0.02 | 2.84 (1.30–6.18) | 0.01 | 2.05 (0.98–4.29) | 0.06 |
| Model fit |  | C-index = 0.70, AIC = 3439.7 |  | C-index = 0.74, AIC = 3368.9 |  | C-index = 0.76, AIC = 3347.7 |  |

**Table 4.** Validation and sensitivity analyses for extent of resection models. AIC = Akaike Information Criterion; C-index = Harrell’s Concordance Index; CI = confidence interval

|  |  |  | Simpson |  | ML-assisted radiology |  | Log-volumetric |  |
| --- | --- | --- | --- | --- | --- | --- | --- | --- |
|  |  |  | Corrected C (CI) | AIC | Corrected C (CI) | AIC | Corrected C (CI) | AIC |
| **Validation** |  |  |  |  |  |  |  |  |
| Bootstrap internal validation |  |  | 0.690 (0.680–0.700) | 3403 | 0.732 (0.725–0.741) | 3361 | 0.747 (0.739–0.756) | 3331 |
| **Sensitivity analyses** |  |  |  |  |  |  |  |  |
| Model performance by location | Convexity | n = 234<br>recurrence = 49 | 0.683 (0.652–0.730) | 472 | 0.716 (0.685–0.759) | 470 | 0.730 (0.699 - 0.775) | 465 |
|  | Parafalcine - parasagittal | n = 230<br>recurrence = 73 | 0.686 (0.656–0.711) | 724 | 0.752 (0.728–0.771) | 698 | 0.763 (0.741–0.782) | 691 |
|  | Skull base | n = 380<br>recurrence = 133 | 0.645 (0.631–0.666) | 1451 | 0.693 (0.682–0.713) | 1430 | 0.715 (0.703–0.735) | 1415 |
| Model performance after adjustment for scan timing |  |  | 0.683 (0.677–0.700) | 2791 | 0.728 (0.721–0.742) | 2748 | 0.744 (0.736–0.757) | 2722 |

We ran two sensitivity analyses. In the first, each multivariate Cox model was restricted to a specific tumour location (e.g. convexity) to assess subgroup performance. The overall ranking of EOR models was preserved across convexity, parafalcine–parasagittal, and skull-base tumours (Table 3). In each subgroup, the log-volumetric model showed the highest corrected discrimination and lowest AIC, ML-assisted radiology performed intermediately, and Simpson grade performed least well. This pattern was most marked in parafalcine–parasagittal and skull-base tumours. The second sensitivity analysis evaluated model performance after additional adjustment for post-operative scan timing. Because some patients had more than one scan within 6 months yielding the same radiological grade, the choice of scan used to derive this timing variable was also varied by permutation in this analysis.

### Nested model comparison

Residual tumour volume provided the greatest incremental prognostic value over the base model without any predictor variables, whereas addition of radiological or Simpson grading to a volumetric model yielded minimal further gain (Supplementary Table 1). Notably, the most parsimonious model with the joint highest C-index was fitted with log-volumetry *and* Simpson grade (corrected C-index 0.75 (0.74-0.76), AIC 2938), possibly reflecting intra-operative information not fully encoded by measured volume alone.

## Discussion

### Summary of results

In this multicentre observational study of cranial meningioma surgery, we compared three competing representations of post-operative extent of resection: surgeon-reported Simpson grade, an ML-assisted radiological grading framework derived from post-operative MRI reports, and semi-automated volumetric measurement of residual tumour burden. A number of key findings emerged. First, concordance between operative and radiological classification differed materially by anatomical compartment, with poorer agreement in skull-base and parasagittal–parafalcine tumours than in more accessible locations. Second, although all three measures were associated with recurrence, increasingly objective measures of residual disease yielded progressively greater prognostic discrimination. Across multiple validation and sensitivity analyses, the log-volumetric model performed best overall, ML-assisted radiological grading performed intermediately, and Simpson grading performed least well. Collectively, these data support a shift in meningioma management away from reliance on intra-operative grading alone toward structured radiological and, where feasible, volumetric assessment of the post-operative residual disease state.

### Interpretation and context

The historical importance of Simpson grading is unquestionable, but its limitations are increasingly apparent in modern neurosurgical practice. Simpson grade remains attractive because it is simple, familiar, heavily embedded in the meningioma literature and still has reliable prognostic utility ^5^. However, it is fundamentally an intra-operative, subjective construct, developed before the routine availability of modern microscopy, neuronavigation, high-resolution MRI, and quantitative post-operative imaging ^16^. Here in the present study, Simpson grading retained prognostic value, but its performance was consistently inferior to both ML-assisted radiology and volumetric residual measurement. This is explained, at least in part, by the fact that the Simpson grade does not directly quantify the amount of residual tumour in contrast to radiological and volumetric approaches.

Concordance analyses provide a plausible explanation for the hierarchy of results observed here. Previous studies have shown that surgeon-reported Simpson grade and post-operative MRI-based classification are related but imperfectly aligned; in 41 meningioma cases, Slot et al. reported absolute agreement of 76% within 72 hours and 78% at 3 months ^17^. Our results are broadly concordant with these findings, but add the important observation that disagreement is anatomically patterned rather than uniform. Lower agreement was seen in falcine, parasagittal, and skull-base tumours than in more surgically accessible lesions. This is consistent with prior evidence that the prognostic utility of Simpson grading varies by dural compartment ^18^, and that EOR is likely to be surgically overestimated when associated with skull-base or sinus infiltration ^2^. In these more complex anatomical settings, operative radicality is frequently limited by narrow surgical corridors, and adjacency to critical neurovascular structures, increasing the likelihood that intra-operative grading will diverge from post-operative imaging-based assessment.

In our work, quantitative residual tumour volume appears to be the most informative representation of meningioma recurrence post-operatively. This finding was maintained after adjustment for tumour location, scan timing (Table 3) and for both short- and longer-term recurrence windows (Supplementary Figure 1). This strongly aligns with previous work by Spille and colleagues in a single centre study of 423 patients, who showed that post-operative tumour volume was independently associated with recurrence and that this recurrence increases with larger residual tumour volume (RTV). In contrast however, Marteri et al. and Brokinkel et al. both failed to demonstrate RTV as a risk factor for recurrence ^19,20^. These conflicting findings may be attributable to sample size, subgrouping and other methodological differences across studies. Our multicentre comparison helps resolve this uncertainty by showing that, when assessed directly against operative and radiological measures, volumetric residual burden provides the most informative prognostic characterisation of post-operative disease, with this finding proving robust across internal validation and sensitivity analyses.

Our findings do not imply that volumetry should replace all other approaches in routine practice. Simpson grade may still contribute complementary information, as shown by the small incremental gain in model fit and parsimony when it was added to the multivariate log-volumetric model (Supplementary Table 1). In addition, imaging-based assessment is vulnerable to confounding from concurrent infection and suboptimal post-operative scan timing, both of which can complicate the distinction between residual tumour and treatment-related enhancement ^6^. Finally, the clinical scalability of volumetry remains constrained by the lack of automated residual-segmentation methods that are sufficiently robust for routine use. Radiological grading consistently outperformed Simpson grade and approached the performance of the volumetric model, particularly in the earlier follow-up period. Prior imaging-pathology studies suggest that post-operative enhancement should not be interpreted uniformly: thin, smooth, continuous dural enhancement is often reactive, whereas thickened, nodular, discontinuous, or tumour-indistinguishable enhancement is more likely to represent residual disease ^7,10^. The value of a structured radiological framework therefore lies in distinguishing likely post-operative change from imaging features that more plausibly reflect biologically meaningful residual tumour.

### Limitations and strengths

Beyond the subjective challenge of distinguishing true residual tumour from reactive post-operative enhancement on MRI, other limitations should be acknowledged. The study is retrospective and relies on routine operative, radiological, and follow-up data, with the attendant risks of inconsistent reporting language, and residual confounding. Variation in post-operative imaging windows will also have introduced some heterogeneity in residual-disease detection, even though sensitivity analyses did not alter the ranking of the competing models. Follow-up was shorter than ideal for a tumour with a long natural history, so the present models are likely to be most informative within the observed follow-up period rather than for very late recurrence. Adjustment for site does not constitute external validation, and transportability remains to be established in independent datasets. Moreover, radiological recurrence does not in itself mandate further intervention, although it often marks an important point of clinical reappraisal and triage.

These limitations are offset by important strengths. This is among the larger multicentre studies to directly compare operative, radiological, and volumetric representations of meningioma EOR against recurrence outcomes. Importantly, this is primarily a characterisation study rather than a definitive prediction-model paper: where the purpose was to determine which representation of post-operative residual disease is most informative, and how these frameworks behave across anatomical subgroups and follow-up time. Within that context, the observed discrimination of the best-performing models was encouraging and comparable against broader multi-modal approaches ^21^. This supports the validity of the present framework while also highlighting substantial scope for improvement through integration of radiomic, molecular, and histopathological data.

## Supporting information

Supplementary Results

Risk Calculator

## Data Availability

Data for this study is available on reasonable request providing ethical permissions have been met

## Conclusion

In conclusion, EOR after meningioma surgery is better captured by objective post-operative residual disease measures than by Simpson grading alone. Although Simpson grade retains prognostic value, ML-assisted radiological grading provides stronger and more reproducible recurrence discrimination, and volumetric residual estimation performs best overall. These findings support a shift toward structured radiological and volumetric assessment of post-operative meningioma state, particularly in anatomically complex tumours where operative grading is most likely to diverge from radiologically identifiable residual disease.

## Supplementary Legends

**Supplementary Figure 1. Time-dependent area under the curve (AUC) for recurrence prediction**

**Supplementary Figure 2. Calibration of extent of resection models for 3-year and 5-year durations**.

**Supplementary Table 1. Nested model performance with addition of predictor variables**. Base variables include age, sex, WHO, tumour location and neurosurgical centre. AIC = Akaike Information Criterion; C-index = Harrell’s Concordance Index; CI = confidence interval; RG = ML-assisted radiology grade; SG = Simpson grade; Vol = log-volumetric

