## Supplementary Methods for "Redefining Extent Of Resection After Meningioma Surgery: a Multicentre Observational Machine Learning Analysis Comparing Simpson, Radiological and Volumetric Grading"

### **ML-assisted radiological grading**

Because post-operative MRI reports were numerous and linguistically heterogeneous across centres, we used a weakly supervised natural language processing approach primarily as an aid to lexicon optimisation. The main purpose of the model was not simply to predict grade, but to identify, in a data-driven manner, the words, phrases, and adjacent synonyms most strongly associated with each radiological residual-disease category, thereby helping harmonise terminology across institutions and improve consistency of classification.

Free-text reports were represented using TF–IDF features derived from word- and character-level n-grams. This representation upweights discriminative terms while downweighting common non-specific language, allowing identification of recurrent reporting patterns linked to particular grades. A LinearSVC classifier was trained using manually assigned labels from a subset of reports. Model development was iterative: feature-weight inspection, review of high-confidence predictions, and structured error analysis were used to identify additional synonym candidates, especially where semantically related terms appeared across neighbouring grades or differed between centres. Through 5 iterations, candidate terms were then incorporated into the evolving radiological grading lexicon and re-evaluated against the manually labelled data. Blank reports, indeterminate reports, and reports containing terms suggestive of infection or reactive change were assigned NA.

Model performance was evaluated to confirm that the learned language patterns were clinically coherent and useful for report classification. In the final analysis, post-operative reports were combined and assessed using grouped 5-fold cross-validation, with MRN as the grouping variable to prevent within-patient leakage. Because Grade 5 was extremely sparse, primary performance metrics were calculated across Grades 1–4. The final model achieved an accuracy of 0.672, macro F1 of 0.669, and weighted F1 of 0.673.
