## Supplementary Results for "Redefining Extent Of Resection After Meningioma Surgery: a Multicentre Observational Machine Learning Analysis Comparing Simpson, Radiological and Volumetric Grading"

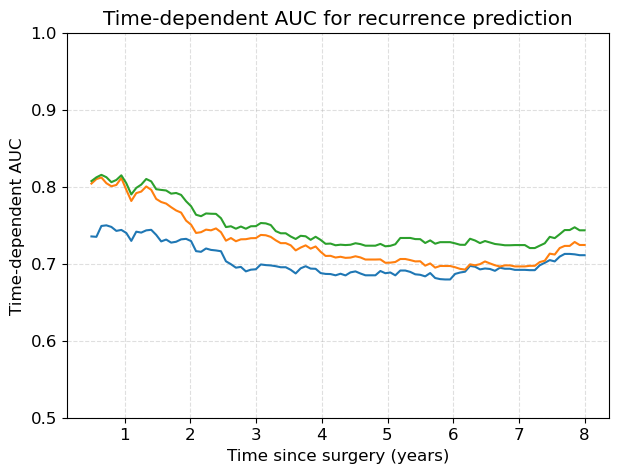

**Supplementary Figure 1. Time-dependent area under the curve (AUC) for recurrence prediction**

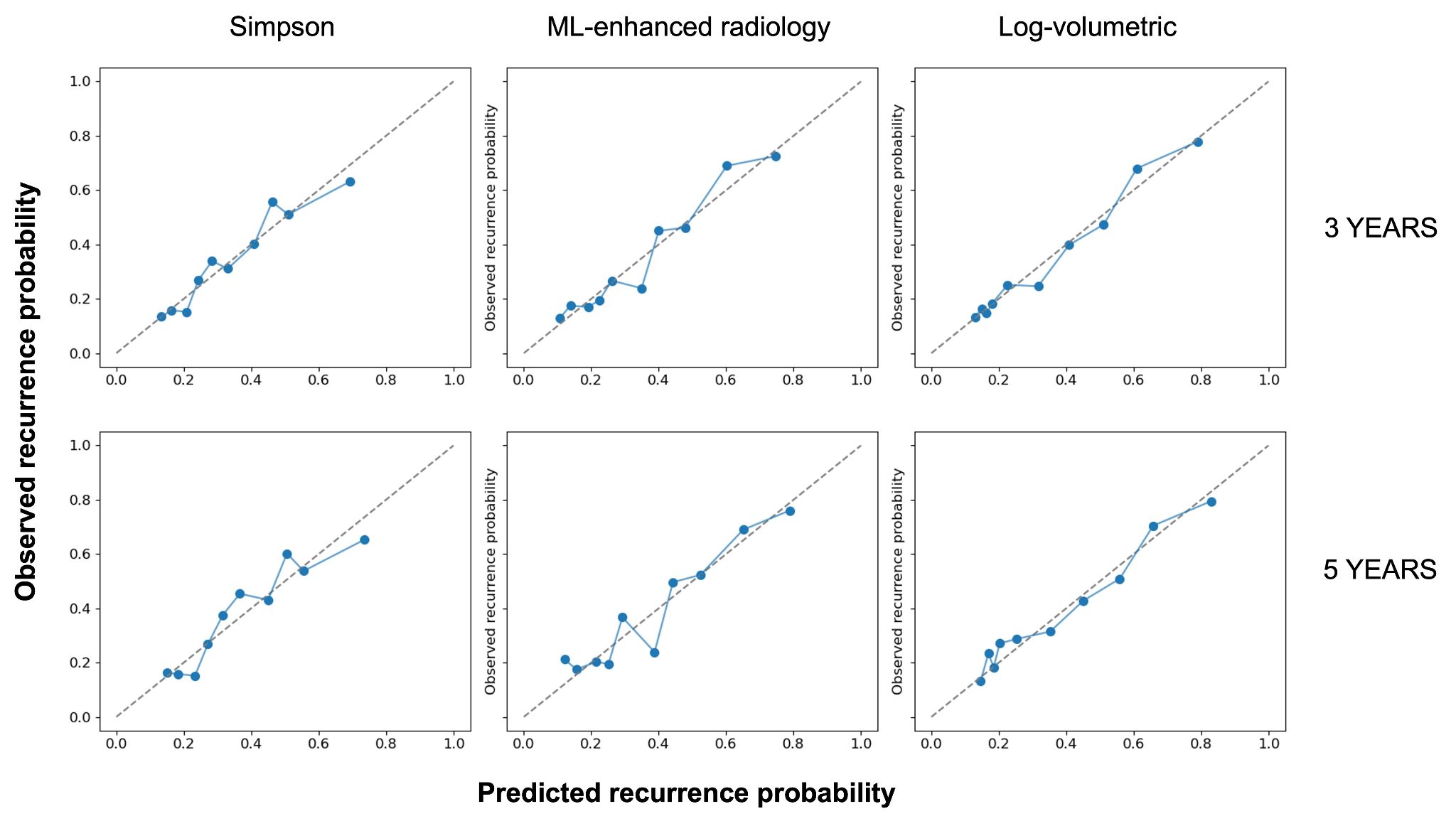

**Supplementary Figure 2. Calibration of extent of resection models for 3-year and 5-year durations.**

|  | **Performance** | |
| --- | --- | --- |
| **Model variables** | **Corrected C-index (95% CI)** | **AIC** |
| **Base** | 0.625 (0.614–0.643) | 3431.262 |
| **Base + SG** | 0.692 (0.681–0.701) | 3376.598 |
| **Base + RG** | 0.733 (0.725–0.740) | 3335.767 |
| **Base + Vol** | 0.748 (0.739–0.754) | 3307.205 |
| **Base + SG + RG** | 0.737 (0.728–0.744) | 3328.373 |
| **Base + SG + Vol** | 0.750 (0.741–0.757) | 3305.932 |
| **Base + RG + Vol** | 0.747 (0.739–0.754) | 3308.429 |
| **Base + SG + RG + Vol** | 0.749 (0.740–0.757) | 3307.444 |

**Supplementary Table 1. Nested model performance with addition of predictor variables.** Base variables include age, sex, WHO, tumour location and neurosurgical centre. AIC = Akaike Information Criterion**;** C-index = Harrell’s Concordance Index; CI = confidence interval; RG = ML-assisted radiology grade; SG = Simpson grade; Vol = log-volumetric
